# Investigation of ventilation conditions associated with CO_2_ concentration changes in ultrasonographic exam room from the perspective of COVID-19 infection control

**DOI:** 10.1101/2021.03.01.21252598

**Authors:** Roka N Matsubayashi, Shino Harada, Mitsuhiro Tominaga

## Abstract

**Objectives:** Ventilation is an important factor in preventing COVID-19 infection. To clarify the state of ventilation in ultrasonic exam rooms, as an index of ventilation rate, the carbon dioxide (CO2) concentration in our exam rooms was measured.

**Methods:** We measured the CO_2_ concentration in each exam room before the examination and 0–15 minutes after end of the exam.

The subjects were 70 cases (abdomen: 24, breast: 16, neck: 16, and musculoskeletal: 14). In infant cases, one parent accompanied the patient during the examination.

**Results:** The highest CO_2_ concentration was 2261 ppm, observed after the breast examination. In all cases, the CO_2_ concentration in the exam room was highest immediately after the examination or two minutes after. Almost all cases had recovered to within 120% of the pre-examination CO_2_ concentrations within 15 minutes after the examination. The average CO_2_ concentration after ultrasonography was significantly higher for breast examinations than others.

**Conclusions:** Even in a hospital with modern ventilation equipment, the CO_2_ concentration in the ultrasound room was high after the exam and it takes 15 minutes to recover to the pre-exam state. Care must be taken to ensure adequate ventilation in ultrasonographic facilities.

## Introduction

The COVID-19 pandemic is ongoing worldwide, including in Japan. At the outbreak’s beginning in Japan, many factors regarding the transmission route were unclear. However, since early March 2020, we have taken prompt measures in consideration of the probable airborne transmission route. Infection control within the Ultrasound Department is important to prevent hospital-related transmission of SARS-CoV-2.

The environmental factors that increase the risk of spreading SARS-CoV-2 are now becoming clear, and knowledge about infection from asymptomatic individuals has been accumulated. Initially, close contact and respiratory droplets were suggested as the main transmission routes of SARS-CoV-2 [1].

Several studies have provided evidence for airborne transmission of viruses, showing that closed, crowded, and poorly ventilated environments contribute to viral transmission [2,3,4]. Recently, airborne transmission has been pointed out as an important pathway for the spread of SARS-CoV-2 [5,6,7,8]. Many research results have indicated that environmental ventilation is an important factor in protection against the spread of COVID-19 [9,10].

Although ventilation equipment is generally designed according to the standards of modern hospitals, complex arrangements and small exam rooms can result in inadequate ventilation in practice. Considering this situation, the air quality of indoor environments should be emphasized [10].

To prevent nosocomial infection, it is particularly important to understand the ventilation situation in hospitals and to implement countermeasures against the spread of COVID-19.

Therefore, we measured carbon dioxide (CO_2_) concentrations in each exam room to assess the effect of subjects’ respiration on room air quality.

Because the examiner was present during the ultrasound examinations, the CO_2_ concentration in each room reflects the respiration of both the subject and the examiner. Thus, we also measured the CO_2_ concentration in the exam room when only the examiner was present for 10 minutes and compared the result to the situation when the subject was also present.

Furthermore, in cases of infants and children aged less than 10 years, a single attendant was present during the examinations, and the observed concentration values are attributable to the respirations of all individuals present.

## Materials and Methods

In this study, we measured the CO2 concentration in the exam rooms in the absence of the patient. No information that identifies the individual patient was used.

Therefore, the Institutional Review Board of the National Hospital Organization Kyushu Medical Center determined that no ethical review was necessary because the study did not involve human subjects.

Our ultrasound center consists of 9 small exam rooms, which are separated by fixed partitions with little space above the partitions (Fig. 1).

**Fig. 1.**
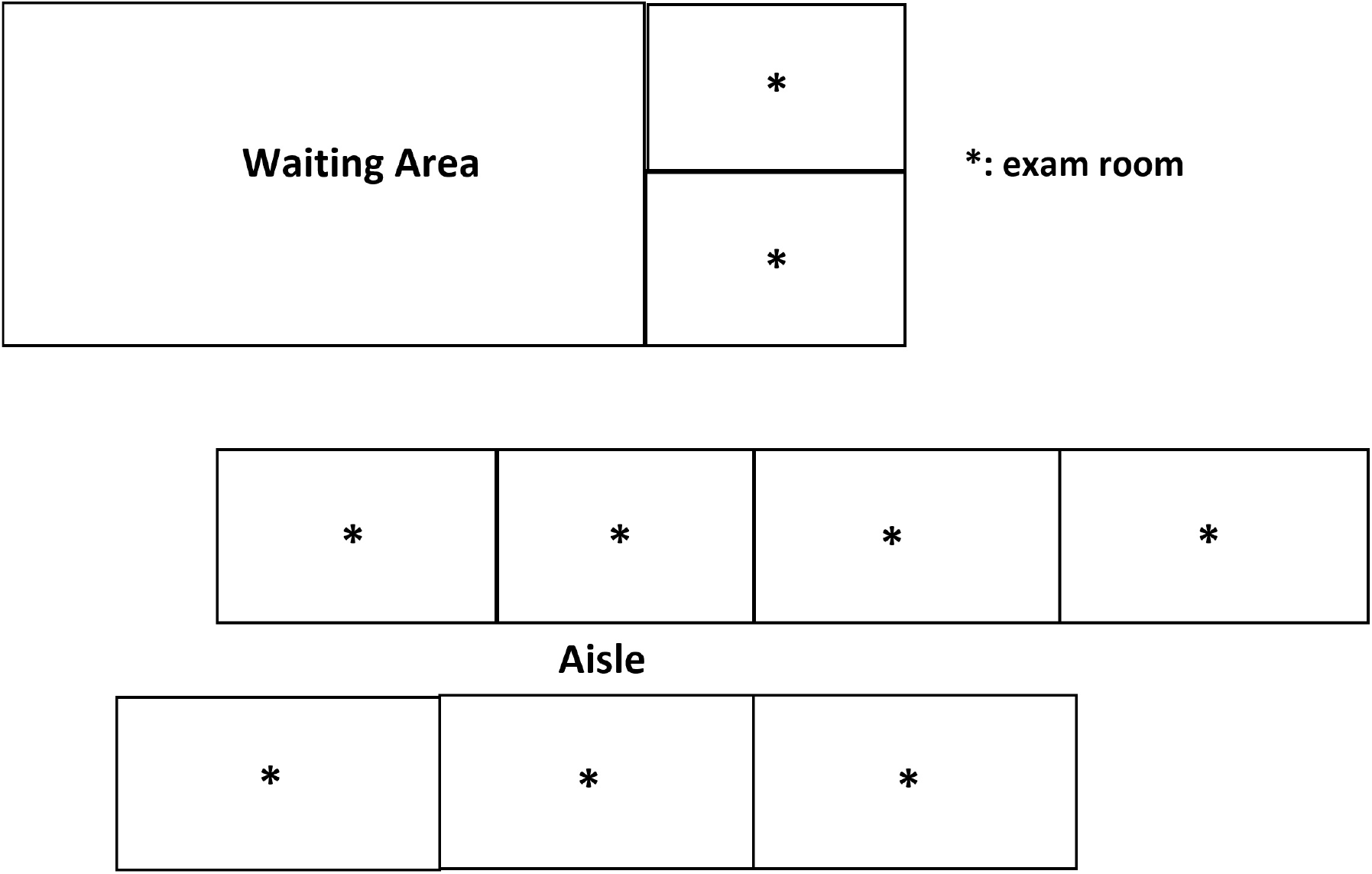
Our ultrasound center consists of 9 small exam rooms, which are separated by fixed partitions with little space above the partitions. Each exam room had an area of 12–15 m^2^.

Each exam room is equipped with air intake and exhaust vents connected to the central ventilation system that was originally installed in each room, which meets the ventilation standards of the Japanese Building Code.

All of these exam rooms and the hospital’s facilities meet Japanese legal environmental standards; however, no evaluation of individual exam room air quality was conducted under actual clinical conditions. The subjects were 70 cases (abdomen: 24, breast: 16, neck: 16, and musculoskeletal (MSK): 14). In the cases of infants and children (abdomen: 6 and musculoskeletal: 6), one attendant accompanied the patient during the examination.

## Methods

In this study, we investigated the CO_2_ concentrations in each exam room before and after the ultrasound examinations. Changes in CO_2_ concentration over time were investigated at multiple time points after completion of the exams.

Measurements were conducted using a multifunctional portable air quality tester (WYZXR Air Quality Monitor, Multifunctional Indoor Pollution Detector Meter). The air quality tester was positioned in the flat part of the ultrasound machine, just between the subject and the examiner (Fig. 2).

**Fig. 2.**
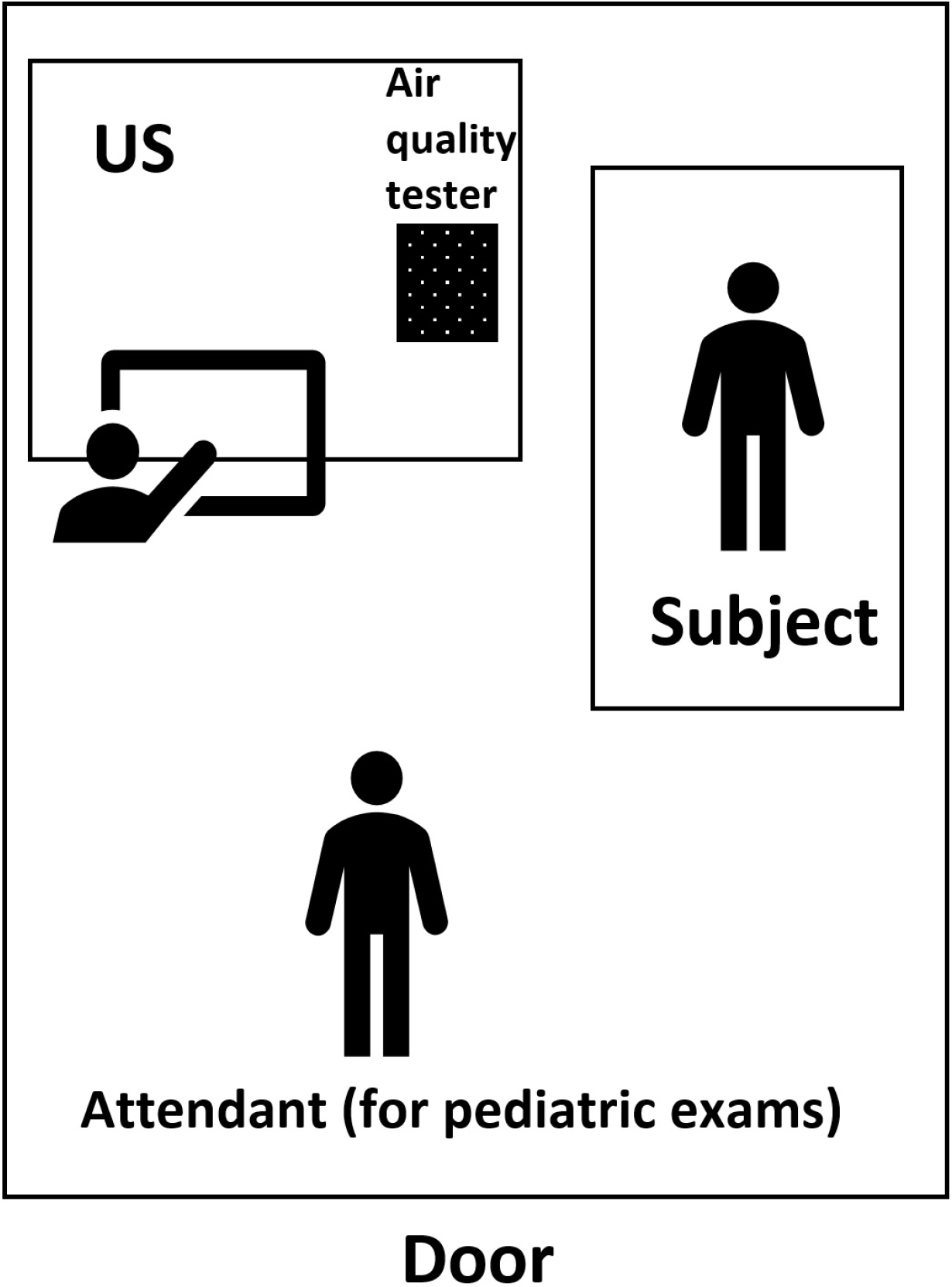
The schematic diagram of CO2 measurement in each laboratory. The air quality tester was positioned in the flat part of the ultrasound machine, just between the subject and the examiner. The height of the air quality tester was 120 cm from the floor.

The height of the air quality tester was 120 cm from the floor, the room temperature was 22–24°C and the humidity was 40%–45%. Each exam room had an area of 12–15 m^2^. All of the exam room doors were kept open and ventilated when ultrasound examinations were not being performed.

After the CO_2_ concentration was measured before each examination, the ultrasound examination was performed with the door closed. After the examination, the CO_2_ concentration was measured at 0–15 minutes (0, 2, 4, 6, 10, and 15 minutes) after the end of the exam. One minute after the end of the examination, the door of the exam room was opened to allow air exchange with the aisle.

For statistical analysis, Bonferroni/Dunn multiple comparison tests were used, with a significance level of p<0.005.

## Results

The average duration of each examination was 10.3±5.6 minutes (3–26 minutes). There were no differences in examination time between body sites.

The highest observed CO_2_ concentration was 2261 ppm, which was observed after breast examination. The CO_2_ concentration was significantly higher in breast examinations than other examinations at all time points after the examination (Table 1)

**Table 1.** Comparison of CO_2_ concentration values (ppm)

|  | Mean | Maximum | Time after examination |  |  |  |  |  |  |
| --- | --- | --- | --- | --- | --- | --- | --- | --- | --- |
|  |  |  | Pre-exam | 0 min | 2 min | 4 min | 6 min | 10 min | 15 min |
| **Abd (n=24)** | 904.6±17 | 1323.5±32 | 544.9±8 | 999.3 | 1291. | 1026. | 878.7 | 685.0±11 | 546.7 |
|  | 9.9 | 8.5 | 7.8 | ±207. | 6±377 | 2±328 | ±262. | 9.4 | ±52.1 |
|  |  |  |  | 2 | .4 | .4 | 2 |  |  |
| **Breast (n=16)** | 1304±113 | 1881.1±18 | 612.3±1 | 1724. | 1827. | 1479. | 1137. | 918.8±16 | 736.8 |
|  | .5 | 1.3 | 29.6 | 9±14 | 3±211 | 2±272 | 4±112 | 3.2 | ±202. |
|  |  |  |  | 9.0 | .2 | .9 | .7 |  | 9 |
| **MSK (n=14)** | 1001.6±1 | 1497.7±18 | 644.6±1 | 1474. | 1283. | 977.6 | 842.6 | 760.1±15 | 669.4 |
|  | 33.0 | 9.9 | 70.4 | 9±19 | 6±163 | ±114. | ±158. | 5.9 | ±150. |
|  |  |  |  | 9.2 | .8 | 6 | 1 |  | 5 |
| **Neck (n=16)** | 678.5±94. | 848.7±166 | 567.5±6 | 698.4 | 762.6 | 748.4 | 711.9 | 600.3±11 | 550.2 |
|  | 6 | .9 | 0.8 | ±229. | ±154. | ±104. | ±130. | 0.4 | ±107. |
|  |  |  |  | 8 | 9 | 6 | 9 |  | 7 |
| **Examiner (n=1)** | 526 | 553 | 533 | 553 | 518 | 539 | 531 | 501 | 507 |
| *p-value* |  |  |  |  |  |  |  |  |  |
| **Abd/breast** | <.0001 | <.0001 | .0725 | <.000 | <.000 | <.000 | <.000 | <.0001 | <.000 |
|  |  |  |  | 1 | 1 | 1 | 1 |  | 1 |
| **Abd/MSK** | .0439 | .0360 | .0117 | .<.00 | .8290 | .5555 | .5725 | .1057 | .0074 |
|  |  |  |  | 01 |  |  |  |  |  |
| **Abd/neck** | <.0001 | <.0001 | .5425 | <.000 | <.000 | .0008 | .0080 | .0585 | .9343 |
|  |  |  |  | 1 | 1 |  |  |  |  |
| **Abd/exami** | .0103 | .0027 | .9189 | .0320 | .0058 | .0547 | .0758 | .1906 | .7692 |
| **ner** |  |  |  |  |  |  |  |  |  |
| **Breast/MS** | <.0001 | <.0001 | .4424 | .0011 | <.000 | <.000 | <.000 | .0022 | .1670 |
| **K** |  |  |  |  | 1 | 1 | 1 |  |  |
| **Breast/nec** | <.0001 | <.0001 | .2721 | <.000 | <.000 | <.000 | <.000 | <.0001 | .0002 |
| **k** |  |  |  | 1 | 1 | 1 | 1 |  |  |
| **Breast/exa** | .<.0001 | <.0001 | .5035 | <.000 | <.000 | .0004 | .0027 | .0041 | .0957 |
| **miner** |  |  |  | 1 | 1 |  |  |  |  |
| **MSK/neck** | <.0001 | <.0001 | .0699 | <.000 | <.000 | .0126 | .0628 | .0021 | .0162 |
|  |  |  |  | 1 | 1 |  |  |  |  |
| **MSK/exam** | .0017 | .0003 | .3491 | <.000 | .0070 | .0872 | .1156 | .0707 | .2387 |
| **iner** |  |  |  | 1 |  |  |  |  |  |
| **Neck/exami** | .2960 | .2400 | .7706 | .4821 | .3747 | .4083 | .3562 | .4824 | .7518 |
| **ner** |  |  |  |  |  |  |  |  |  |

*MSK* musculoskeletal
*Abd* abdomen
*p*<.005

(Fig. 3).

**Fig. 3.**
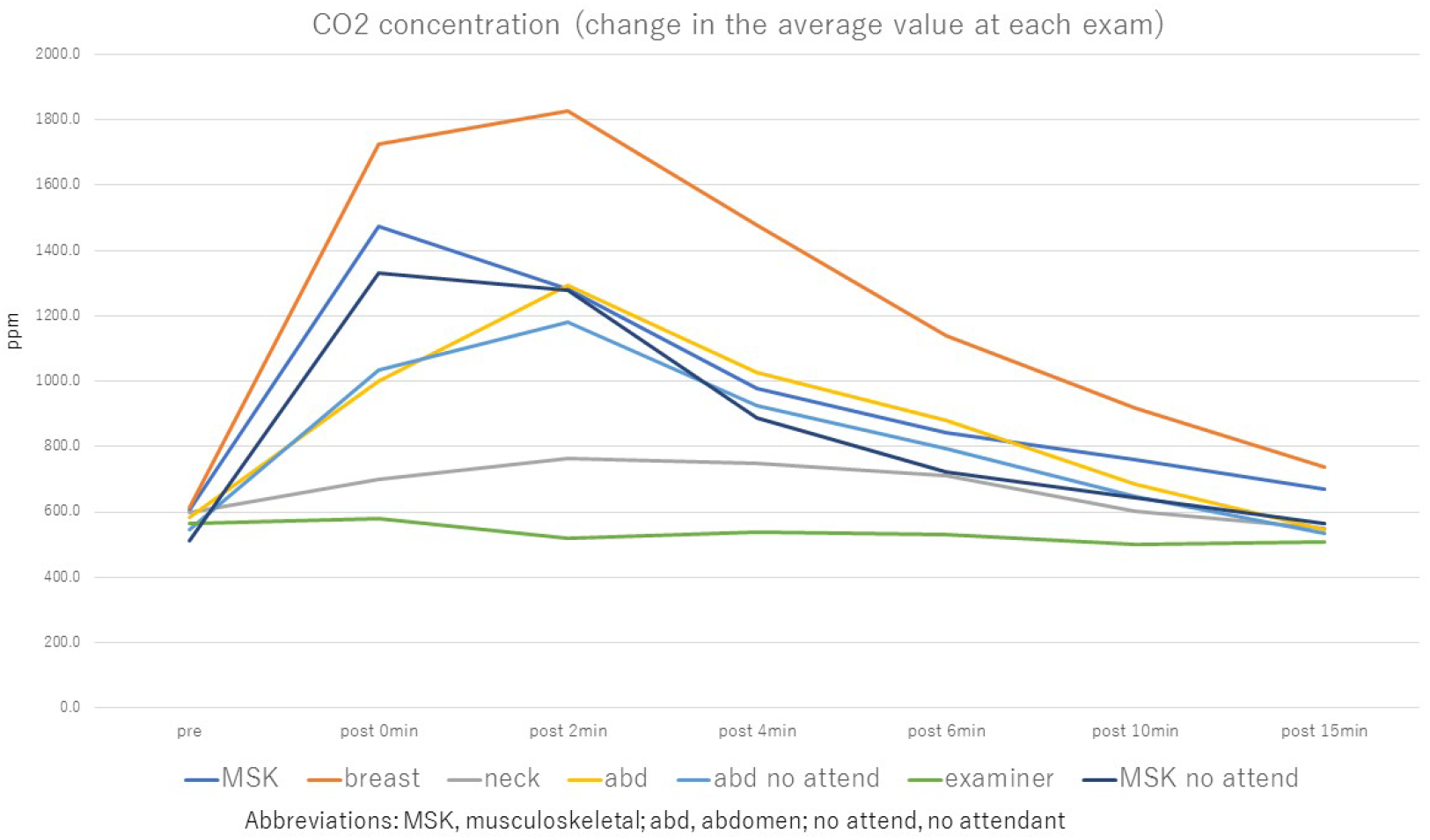
Change in the average CO2 concentration over time for each test site. The CO_2_ concentration was significantly higher in breast examinations than other examinations at all time points after the examination. *MSK, musculoskeletal; abd, abdomen; no attend, no attendant*

In half of breast examination cases, the highest CO_2_ value was shown 2 minutes after the examination. All six infant abdominal cases had the highest observed CO_2_ concentration 2 minutes after the examination. In other cases, the highest CO_2_ concentration was observed immediately or 2 minutes after the examination, and the value then decreased over time.

A comparison of mean and maximum CO_2_ concentrations during post-test measurements among the sites showed that the breast had the highest values (p<0.0001) (Table 1) (Fig. 4).

**Fig. 4.**
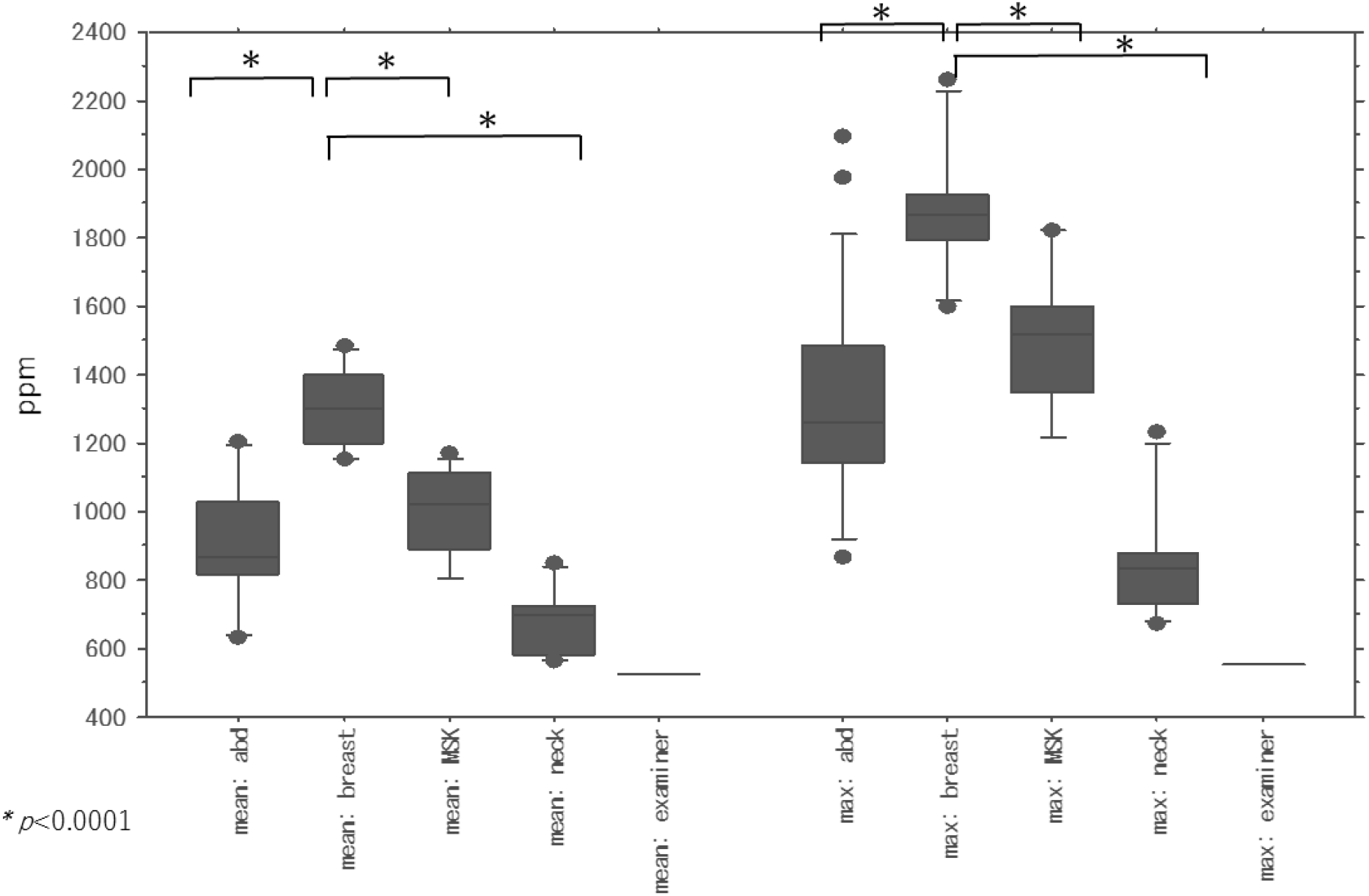
A comparison of mean and maximum CO_2_ concentrations during post-test measurements among the sites showed that the breast had the highest values (\**p*<0.0001). *MSK, musculoskeletal; abd, abdomen; no attend, no attendant*

Except in the case of the single examiner alone in the exam room, the lowest mean and maximum CO_2_ concentrations were significantly lower in neck examinations.

Both mean and maximum concentrations were significantly higher in the presence of an attendant than with the subject alone (mean concentration: p=0.0017, maximum concentration: p=0.007) (Fig. 5).

**Fig. 5.**
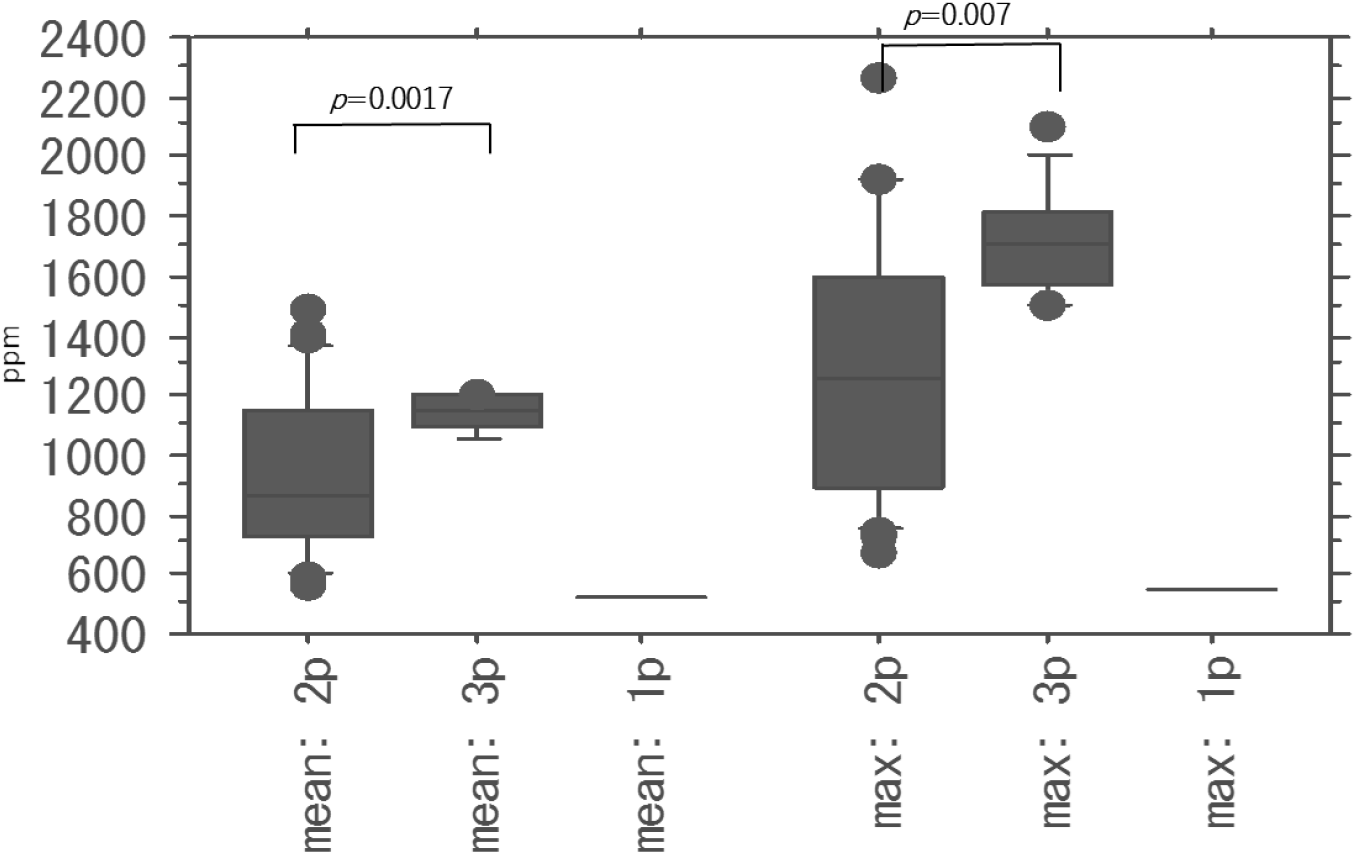
Both mean and maximum concentrations were significantly higher in the presence of an attendant than with the subject alone (mean concentration: p=0.0017, maximum concentration: p=0.007). *1p: one person (examiner alone), 2p: two persons (subject and examiner), 3p: three persons (subject, examiner, and attendant)*

We compared the ratio of the CO_2_ concentration at each time point after the end of the test to the CO_2_ concentration before the start of the test. The mean CO_2_ concentration at all sites had recovered to within 120% of the pre-test CO_2_ concentration at 15 minutes after the end of the test, but the value was higher in the case of the breast at all time points, and the concentration was still significantly higher 15 minutes after breast than neck examinations (Table 2)

**Table 2.**
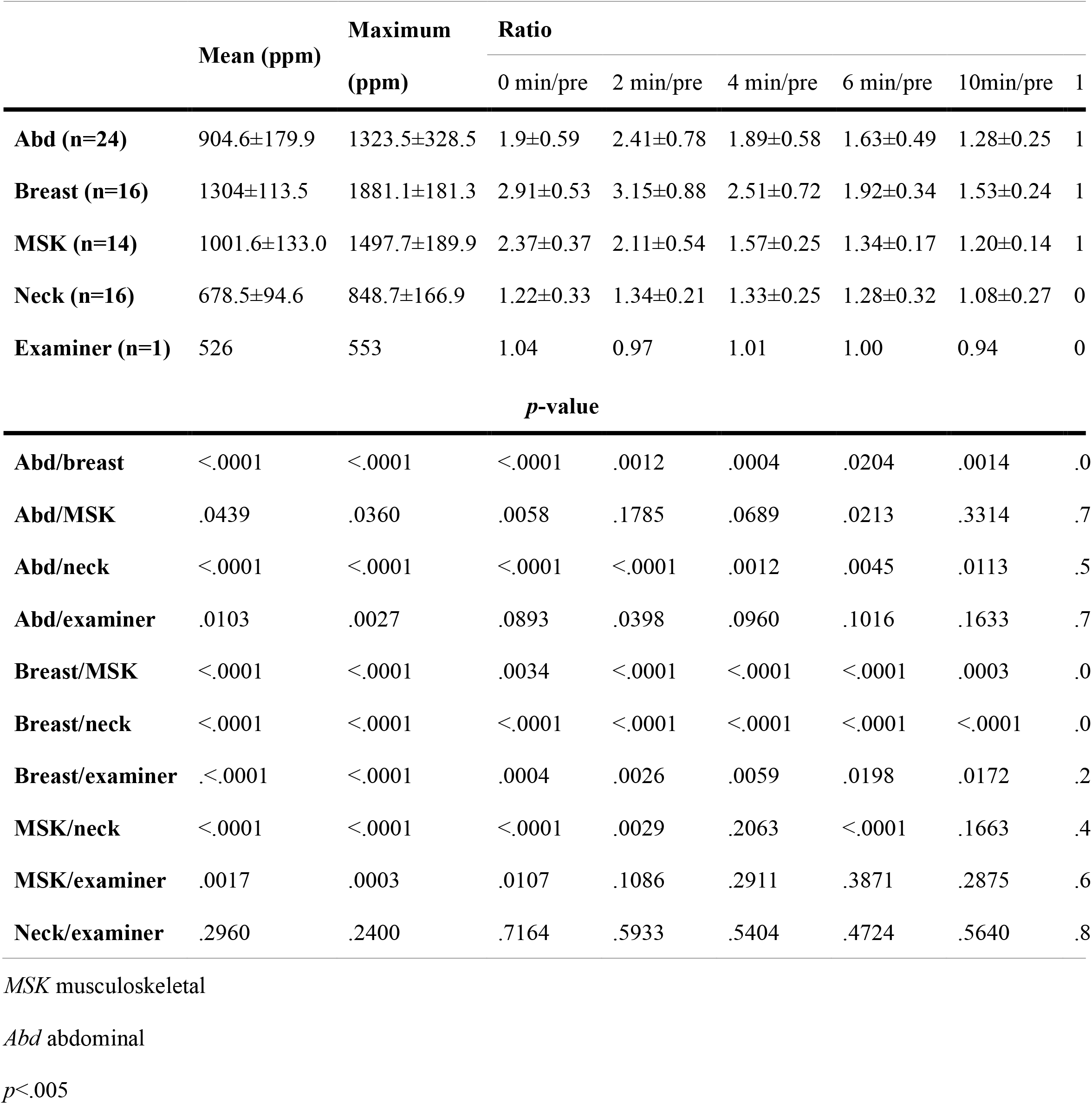
Comparison of CO_2_ concentrations in each time phase.

(Fig.6).

**Fig. 6.**
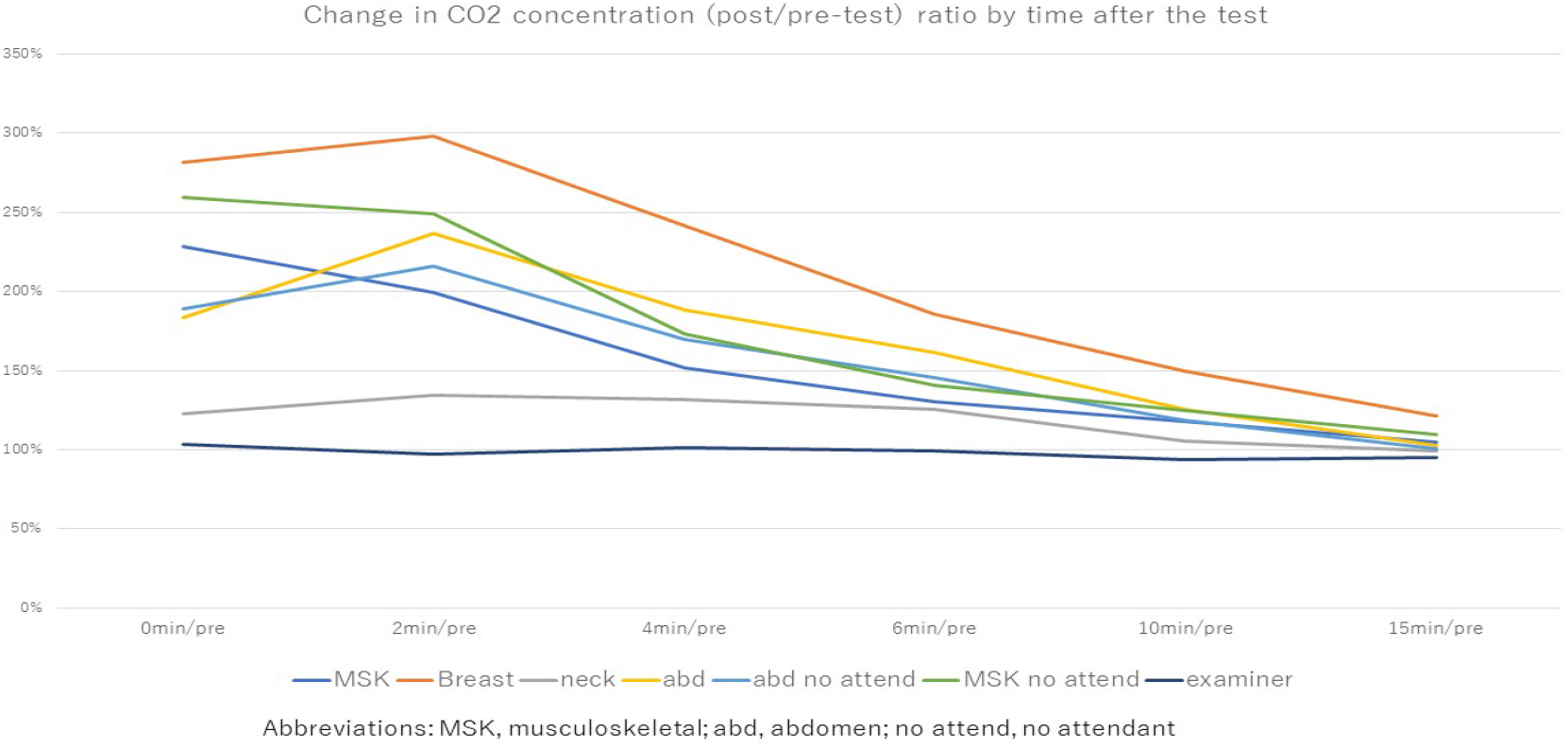
Comparison of the ratio of the CO_2_ concentration at each time point after the end of the test to the CO_2_ concentration before the start of the test. The mean CO_2_ concentration at all sites had recovered to within ±20% of the pre-test CO_2_ concentration at 15 minutes after the end of the test, but the value was higher in the case of the breast at all time points, and the concentration was still significantly higher 15 minutes after breast than neck examinations. *1p: one person (examiner alone), 2p: two persons (subject and examiner), 3p: three persons (subject, examiner, and attendant)*

In the case of the examiner being alone in the room, CO_2_ concentrations were unchanged throughout all time phases, with a maximum of 103% of the pre-test value (immediately after the end of the test).

## Discussion

In the present study, we measured the CO_2_ concentration in ultrasound exam rooms with the aim of evaluating the ventilation in ultrasound rooms under real clinical conditions.

The results show that CO_2_ concentrations were elevated by breast examinations and the presence of an attendant in pediatric cases. In contrast, CO_2_ concentrations remained low during neck examinations compared with those of all other areas.

A possible reason for the lack of elevated CO_2_ concentrations is that the subjects did not vocalize during the neck examinations. In contrast, in breast and pediatric cases, the examiner needs to talk to the patient or attendant to understand the lesion’s location and symptoms, possibly influencing the increase in CO_2_ concentration. Examinations of the abdomen, which require repeated inhalation/exhalation during the test, showed lower CO_2_ concentrations than those of the mammary gland, suggesting that CO_2_ concentrations may be elevated by vocalization and speech.

The observed changes in CO_2_ concentration over time suggest that air conditions would not recover to their pre-exam state for about 15 minutes in the absence of forced ventilation, suggesting that mechanical ventilation or a circulator to create air flow would be desirable.

The current study has several limitations: it is a study of a single facility and a specific environment, and more precision-controlled methods are required for a rigorous assessment of air quality.

## Conclusions

We measured CO_2_ concentrations before and after testing in relatively small ultrasound rooms and examined the concentration ratios and changes over time.

Even at a hospital with modern ventilation equipment, the CO_2_ concentration in the ultrasonic exam room rises immediately after the examination, and it takes at least 15 minutes to recover to the approximate concentration observed before the examination. In ultrasonographic facilities, care must be taken to ensure adequate ventilation.

## Data Availability Statement

The data that support the findings of this study are available from the corresponding author, RM, upon reasonable request.

